# The relationships between ongoing COVID-19 lockdown and the financial and mental health experiences of Australian families

**DOI:** 10.1101/2021.08.15.21262087

**Authors:** Anna M. H. Price, Diana Contreras-Suárez, Anna Zhu, Natalie Schreurs, Mary-Anne Measey, Sue Woolfenden, Jade Burley, Hannah Bryson, Daryl Efron, Anthea Rhodes, Sharon Goldfeld

## Abstract

**Objectives:** In 2020, Australia’s successful COVID-19 public health restrictions comprised a national ‘initial lockdown’ (March-May), and ‘ongoing lockdown’ (July-November) for metropolitan Victorian residents only. We evaluated the relationships between ongoing lockdown and family finances and mental health.

**Methods:** In the June and September 2020 Royal Children’s Hospital National Child Health Polls, caregivers of children in Victoria and New South Wales reported: job/income loss; material deprivation (inability to pay for essential items); income-poverty; mental health (Kessler-6); perceived impact on caregiver/child mental health; and caregiver/child coping. Data from N=1207/902 caregivers in June/September were analysed using Difference-in-Difference modelling (New South Wales provided the comparator).

**Results:** During Victoria’s ongoing lockdown, job/income loss increased by 11% (95%CI: 3-18%); Kessler-6 poor mental health by 6% (95%CI: -0.3-12%) and perceived negative mental health impacts by 14% for caregivers (95%CI: 6-23%) and 12% for children (95%CI: 4-20%). Female (versus male) caregivers, metropolitan (versus regional/rural) families, and families with elementary school-aged children (versus pre-/high-school) were most affected.

**Conclusions:** Ongoing lockdown was associated with negative experiences of mental health, employment, and income, but not deprivation or poverty, likely because of government income supplements introduced early in the pandemic. Future lockdowns require planned responses to outbreaks, and evidence-informed financial and mental health supports.

## Introduction

The coronavirus SARS-COV-2 (COVID-19) was first identified in Australia in late January 2020. From March, Australian governments at the federal and state levels implemented a range of public health restrictions, including stay-at-home orders (also known as ‘lockdown’). In 2020, Australia’s lockdown response was among the most stringent internationally.(1) By 31 December, the measures successfully contained infection to an overall incidence rate of 111 cases and 3.5 deaths per 100,000 people.(2) In contrast, other high-income countries with more lenient public health restrictions, such as the United States (US) and United Kingdom (UK), recorded rates of 5895 and 3730 cases per 100,000 people, respectively.(2) Studies of previous pandemics and from the first months of COVID-19 showed that quarantine and isolation could have indirect and negative impacts on household finances and mental health.(3-10) This is particularly the case for families with children, where there are mixed views on the balance of harms versus benefits of lockdown.(6, 10-13) Unlike many high-income countries, Australia’s low incidence of COVID-19 in 2020 made it possible to examine the effects of lockdown mostly independent of the compounding disease impacts, which forms the purpose of this paper.

The evolution of Australia’s COVID-19 public health restrictions is presented in Supp. Figure 1. From 23 March to 1 June 2020, the national ‘initial’ lockdown included mandatory quarantine for returned travellers; travel bans; self-isolation for suspected/confirmed cases; stay-at-home orders; and closure of schools and ‘non-essential’ businesses.(10, 14, 15) Five weeks after initial lockdown eased, a second wave of infections in the state of Victoria rapidly surpassed active cases in the first wave, with the national peak reaching 721 new cases (2.8 per 100,000) in 24 hours. From 8 July-23 November 2020, Victorian residents entered an ‘ongoing’ and more severe lockdown. The public health measures were strictest for metropolitan areas (in the state’s capital city of Melbourne) compared with regional and rural Victoria. Previous lockdown measures were reinstated, the stay-at-home orders were further restricted, a night-time curfew was added, and early childhood education and care providers closed (see Supp. Figure 1). Compliance with Australia’s lockdown measures was driven by state enforcement, through police surveillance and fines.

To protect against the economic fallout of lockdown, the Australian federal government rapidly implemented a suite of short-term financial supports.(16, 17) Shown in Supp. Figure 1, they included an unemployment supplement (‘JobSeeker’) which doubled recipients’ social welfare benefits from $550 to $1,100 a fortnight;(16) a wage supplement for eligible businesses to retain their workforce (‘JobKeeper’);(17) allowing early access to superannuation;(18) and free childcare for working families.(19) Banks and creditors also allowed loan repayments to be deferred for up to 10 months. These social policy changes represent some of the largest (albeit temporary) in Australia’s history. Indeed the JobKeeper and JobSeeker supplements were so significant that, by September 2020, levels of poverty and housing stress in Australia were substantially lower than the levels directly preceding COVID-19.(16)

While these social policies buffered Australians from poverty, global data on COVID-19 show that lockdown has substantial and negative indirect impacts on households. In a review of the global mental health evidence from the first year (to April 2021), Aknin et al., reported a peak in adults’ psychological distress in the early months.(20) While many studies reported a decline to pre-pandemic levels by mid-2020,(20) the authors found that mental health inequities were sustained or exacerbated for adults who were younger, female, child-rearing, or with fewer socioeconomic resources.(20, 21) This is supported by Australian cross-sectional data of 1200 adults repeated weekly from March 2020, which found that mental distress tripled for parents from 8% pre-COVID-19 to 24% during the pandemic.(22) The same survey data showed similar patterns for self-reported financial stress and material deprivation (unable to afford essential items).(22) The latter aligns with the notion of the pandemic being syndemic; that is, synergistically acting with current inequities to exacerbate the negative impacts of social demographics including indicators of adversity.(23)

While the infection and mortality rates of the original COVID-19 strains were lower in children than adults,(24,25) children are more developmentally vulnerable to their socioeconomic environment than adults.(26,27) In Racine and colleagues’ (2021) meta-analysis of 29 studies published in the first year of the pandemic, prevalence estimates for depression and anxiety in children doubled.(28) Previous studies show that the stress and isolation of lockdown can negatively impact children’s mental health for many months,(29) and school closures can compromise children’s educational opportunities for years.(6, 11)

To our knowledge, no studies have investigated the relationships between COVID-19 lockdown and both the financial and mental health experiences of families with children, in the relative absence of disease morbidity and mortality. This evidence can provide insights into the potential impacts of lockdown to help inform economic and population health responses to future public health crises. This study uses the natural experiment that occurred in Australia, whereby the state of Victoria experienced ongoing and more severe lockdown, to address this evidence gap. Data are drawn from the Royal Children’s Hospital (RCH) National Child Health Poll, the only nationally representative survey to measure families’ and children’s experience of the COVID-19 pandemic. The Poll was conducted in June (when the initial lockdown had ended for all Australians) and in September 2020 (when only metropolitan Victorians were in ongoing, stricter lockdown). Data from the neighbouring state of New South Wales (NSW), which experienced only the initial lockdown, provide the comparator. The two states are inherently similar in terms of their population, size, and geographic location.

The specific aims were to (1) describe families’ financial and mental health experiences after the initial lockdown and (2) evaluate the relationship between ongoing lockdown on family finances and mental health (a) overall and (b) by caregiver gender, child age, and geographical location. We hypothesised that the ongoing lockdown would be associated with increased financial hardship and worse mental health.

## Methods

### Design and procedure

The RCH National Child Health Poll comprises periodic cross-sectional surveys of approximately 2000 Australian caregivers of children aged 0-17 years. Data collection is contracted to the Online Research Unit who obtain written informed consent and draw a nationally representative sample of caregivers using stratified random sampling from their panel of over 350,000 adults aged 18 years or older, who live in Australia and have internet access. Panel members have a unique identifying number that means they can only access and complete the Poll once. Only one person per household can join the panel. The field period for each Poll is approximately two weeks. Surveys are administered in sixth grade-equivalent English, which corresponds to the end of elementary school (known as primary school in Australia). Responses are anonymous, and respondents are remunerated with points exchangeable for department store gift vouchers. Surveys reported in this paper were conducted during 15-23 June and 15-29 September 2020 with two different samples. They focused on families’ experience of financial hardship and mental health 3 and 6 months into the COVID-19 pandemic. The RCH Human Research Ethics Committee approved the research (February 2020, #35254).

### Patient and Public Involvement

RCH Polls are informed by previous surveys, which ask caregivers to identify the child health issues of most concern to them and which child health topics should be included in future polls. At the end of each survey, participants were informed of the study website where all research reports are accessible to the public. As each survey is collected from a cross-sectional, population-based online survey of a random sample, respondents were not directly involved in the recruitment or conduct of the study.

### Measures

Both Polls analysed in this study captured demographic information including caregiver and child age and gender, number of children in care, caring for a child with additional health needs (chronic illness, health condition, disability), partner status, Healthcare Card status (identifies low-income), caregiver education level, identifying as Aboriginal or Torres Strait Islander, country of birth, language spoken at home, living in metropolitan/regional/rural areas and postcode.

Family finances were assessed using:

1. Nine items adapted from the CoRonavIruS Health Impact Survey (CRISIS) caregiver version (30): “What changes in employment or income have occurred in your household due to coronavirus/COVID-19?” (response options “yes” versus “no”) including: “job loss by one caregiver”; “job loss by two caregivers”, “difficulty paying bills or for necessities”, “working longer hours”, “filing for unemployment”; “applying for Government assistance”; “reduced work hours”; “reduced total household income” or “none of the above”. A binary variable describing any job loss (by one or two caregivers) or reduction in income due to COVID-19 (versus not) was created to enable comparison with other Australian studies, e.g. (31)
2. Eight items adapted from the Household, Income and Labour Dynamics in Australia (HILDA) Survey Wave 18 Household Questionnaire Material Deprivation Module (32) asking “In the last month, because of money pressure did you miss or put off” (response options: “yes” versus “no”): mortgage or rent repayments; electricity, gas, water bills; food; healthcare; prescription medicines; home or car insurance; mobile phone bills; and internet. Two summary variables were created: (a) a binary “any material deprivation” variable identifying inability to pay for one or more essential items (versus “none”), and (b) a “total material deprivation count” summing the number of essential items where payment was missed or put off (possible range 0-8).
3. Current total household income before tax, categorised into 10 options ranging from “less than $500 p/week” to “more than $3,000 p/week”, plus “prefer not to say”. A binary variable was created to summarise low income (“less than AU$1,000 p/week” versus more) based on Australian thresholds for HealthCare Card eligibility and definitions of income-poverty.(33) Mental health was assessed using:
4. 6 items of the Kessler-6 (K6) assessing caregivers’ self-reported anxiety and depressive symptoms encountered in the last 4 weeks. Scored on a 5-point Likert scale from 1 “none of the time” to 5 “all of the time”. Summarised into (a) a continuous total score, and (b) a binary variable using the established cut-point for the Australian population identifying “poor mental health” (total score 19 or more) versus not (total score 6-18).(34)
5. A 5-point item adapted from UK Young Minds Matter 35) asking “What would you say the impact of COVID-19 has been on your mental health / the mental health of your child?” dichotomised into perceived negative impacts (“small negative impact/large negative impact”) versus perceived positive impacts (“no impact/small positive impact/large positive impact”). Reported by caregivers for (a) themselves and (b) each child.
6. A 4-point study-designed item asking ““Which of the following best describes how you are / your child is managing with life at the moment?” reported by caregivers and dichotomised into “struggling/not coping” versus “coping/thriving”. Reported by caregivers for (a) themselves and (b) each child.

### Analysis preparation

Families living in the Australian states of Victoria and NSW were retained in the analytic sample. For each family, we assigned data from the Australian Bureau of Statistics (ABS) Socio-Economic Indexes for Areas (SEIFA) Index of Relative Disadvantage, a national area level index derived from census data for all individuals living in a postcode, with higher scores indicating greater advantage. Forty-one families (with 63 children) preferred not to report their country of birth, and one family (with two children) were missing SEIFA (see Supp. Figure 2). As country of birth and SEIFA were included as potential confounding variables (controls), the records with missing data were dropped by the regression analyses. Thus, to accurately represent the analytic sample, the small number of records with missing data (2%) were also excluded from the descriptive analyses. Measures were weighted to reduce the effects of non-response and non-coverage and therefore approximate the population distributions of financial and mental health experiences. Weights were derived using the ABS 2016 Census of Population and Housing, Customised Data Report, according to distributions of caregiver age, gender, family structure (sole-caregiving, number of children and any under 5 years), state/territory and SEIFA.

### Analysis

Demographics were described by survey and state using unweighted data. Family finances and mental health experiences (Aim 1) were described by survey and state using weighted proportions for categorical data and mean and standard deviation (SD) for continuous data. The change in family finances and mental health as related to lockdown (Aim 2a) was estimated using Difference-in-Difference analyses implemented as an interaction term between time (September versus June) and treatment group dummy variables (Victoria versus NSW) using linear regression models. The Difference-in-Difference approach is appropriate given the aim and the policy set-up. Ongoing lockdown was introduced only for Victoria and not for NSW. This created a natural experiment that allowed comparison of a group of families who were exposed to ongoing lockdown (Victoria) with an unexposed group (NSW). The Difference-in-Difference estimator compares families’ outcomes before and after the policy implementation.

Given the emerging evidence showing the differential impacts of the pandemic according to family and socioeconomic characteristics,(20-22) the Difference-in-Difference models controlled for demographic variables that were available in the dataset and likely to confound the relationship between lockdown and outcomes. These included: child and caregiver age and gender (male/female); number of children; child with additional health needs (versus not); one-caregiver family (versus not); owns a Health Care Card (versus not); caregiver education (<Year 10/Year 10/Year 12/trade or apprenticeship/certificate or diploma/undergraduate/postgraduate); Aboriginal or Torres Strait Islander (versus not); caregiver born outside of Australia (versus in Australia); Home language other than English (versus speaks English at home); lives regionally or rurally (versus metropolitan); and SEIFA quintile. Caregiver mental health (total K6 score) was included as a control in the regressions for child mental health. All child models included family clustered errors to account for the correlation that exist at this level. These models were repeated for three subgroups (Aim 2b): (i) caregiver gender (female/male); (ii) child age (grouped at 0-4 years (as a proxy for preschool), 5-11 years (a proxy for elementary/primary school), 12-17 years (a proxy for secondary/high school); (iii) and metropolitan/regional/rural location. The latter was used as a proxy for severity of lockdown, noting that metropolitan Victorians endured a more severe and longer lockdown than their regional and rural counterparts but also that the whole state was affected. Subgroup models controlled for the same variables except the grouping variable, which provided the strata for analysis.

We chose to run linear regression models as interpretation is simpler than logistic regression and most of the predicted y-values were bounded within 0-1. To check the robustness of these models, we ran marginal effects probit models for the dichotomous outcomes for the whole sample (not presented), which confirmed the linear regression output. As 293 caregivers preferred not to disclose their income, these analyses should be interpreted with caution. Data were analysed with Stata v17.

## Results

### Sample characteristics

Supp. Figure 2 presents the respondent flowchart for the analytic sample. In June, after initial lockdown ended, 2697 Australian caregivers were invited and 2020 (75%) completed the Poll. Of these, the 1207 families with 1992 children who lived in Victoria (604 caregivers/985 children) or NSW (603 caregivers/1008 children) were included in the analysis. In September, during Victoria’s ongoing lockdown, 1769 caregivers were invited and 1434 (81%) completed the Poll. Of these, the 902 families with 1584 children who lived in Victoria (460 caregivers/786 children) or NSW (442 caregivers/798 children) were included in the analysis.. Table 1 describes the demographic characteristics by survey and state. The SEIFA quintiles suggested strong response bias towards more socio-economically advantaged groups. There were some differences between surveys in characteristics such as the proportion of respondents caring for young children, caregiver gender, sole caregiving, and SEIFA; characteristics that were used to create the sample weights and analytic controls (see Analysis).

**Table 1.** Demographic characteristics (unweighted) of New South Wales and Victorian families surveyed after initial lockdown (June 2020) and during Victoria’s ongoing lockdown (September 2020) in proportions unless specified.

| Demographic characteristics | New South Wales |  | Victoria |  |
| --- | --- | --- | --- | --- |
|  | June | September | June | September |
| **Caregiver** | **N=603** | **N=442** | **N=604** | **N=460** |
| Age (years), *mean (SD)* | 44.4 (9.3) | 41.4 (7.8) | 43.9 (9.1) | 42.2 (8.3) |
| Number of children in care, *mean (SD)* | 1.7 (0.7) | 1.8 (0.8) | 1.6 (0.8) | 1.7 (0.8) |
| Female | 47.3 | 52.3 | 55.4 | 52.3 |
| One-parent family | 25.2 | 20.6 | 22.2 | 17.6 |
| Caregiver to child with additional health needs <sup>a</sup> | 40.3 | 48.2 | 39.2 | 44.1 |
| Health Care Card (identifies low income) | 14.8 | 21.7 | 14.1 | 17.8 |
| Education |  |  |  |  |
| Less than Year 10 | 1.2 | 1.4 | 0.8 | 1.7 |
| Year 10 | 5.5 | 5.7 | 3.2 | 2.6 |
| Year 12 | 8.6 | 8.8 | 12.8 | 12.6 |
| Trade / apprenticeship | 3.8 | 5.7 | 2.5 | 2.8 |
| Certificate / diploma | 19.4 | 22.6 | 21.7 | 18.5 |
| Undergraduate | 38.5 | 32.6 | 37.6 | 41.5 |
| Postgraduate | 23.1 | 23.3 | 21.5 | 20.2 |
| Aboriginal or Torres Strait Islander | 2.8 | 5.2 | 1.5 | 4.6 |
| Born in Australia | 27.4 | 21.5 | 26.7 | 22.8 |
| Home language other than English | 23.2 | 29.0 | 23.2 | 23.0 |
| Lives regionally or rurally (versus metropolitan) | 12.1 | 11.8 | 15.7 | 11.5 |
| SEIFA Index of Social Disadvantage Quintile |  |  |  |  |
| 1 (most disadvantage) | 11.0 | 13.1 | 8.3 | 10.7 |
| 2 | 15.1 | 18.8 | 16.2 | 19.4 |
| 3 | 17.4 | 20.8 | 17.4 | 15.9 |
| 4 | 16.3 | 13.1 | 23.7 | 23.7 |
| 5 (least disadvantage) | 40.3 | 34.2 | 34.4 | 30.4 |
| **Child** | **N=1008** | **N=798** | **N=984** | **N=786** |
| Age (years), *mean (SD)* | 10.2 (4.9) | 9.1 (4.7) | 9.9 (5.0) | 9.5 (4.9) |
| Female | 48.7 | 49.4 | 49.0 | 49.4 |

CI: 95% Confidence Interval. SEIFA: Socio-Economic Indexes for Areas.
<sup>a</sup> Examples of additional health needs include asthma, epilepsy, and diabetes; of disability includes hearing impairment, intellectual disability, and autism.

### Experiences after initial lockdown (Aim 1)

Table 2 describes families’ financial and mental health experiences in June 2020. Table 3 presents the mean differences (MDs) between NSW and Victoria (‘Vic June’ column; see footnote for controls). Twenty-nine percent of NSW caregivers reported job or income loss due to the COVID-19 pandemic, compared with 24% of Victorians (MD=5%; 95% CI: 0.1 to 10%). Tables 2 and 3 show that financial and mental health experiences were otherwise similar between states. Fifteen percent of families reported low income, and one-third reported material deprivation (unable to pay for essential items including mortgage or rent; electricity, gas, water bills; food; healthcare; prescription medicines; home or car insurance; mobile phone bills; and internet). Caregivers averaged one missed payment out of the eight essential items. One in five caregivers reported poor mental health according to the K6; half said the pandemic had negative impacts on their mental health; and one quarter said it had negative impacts on their child’s mental health. Despite these negative experiences, most caregivers and children reported coping.

**Table 2.**
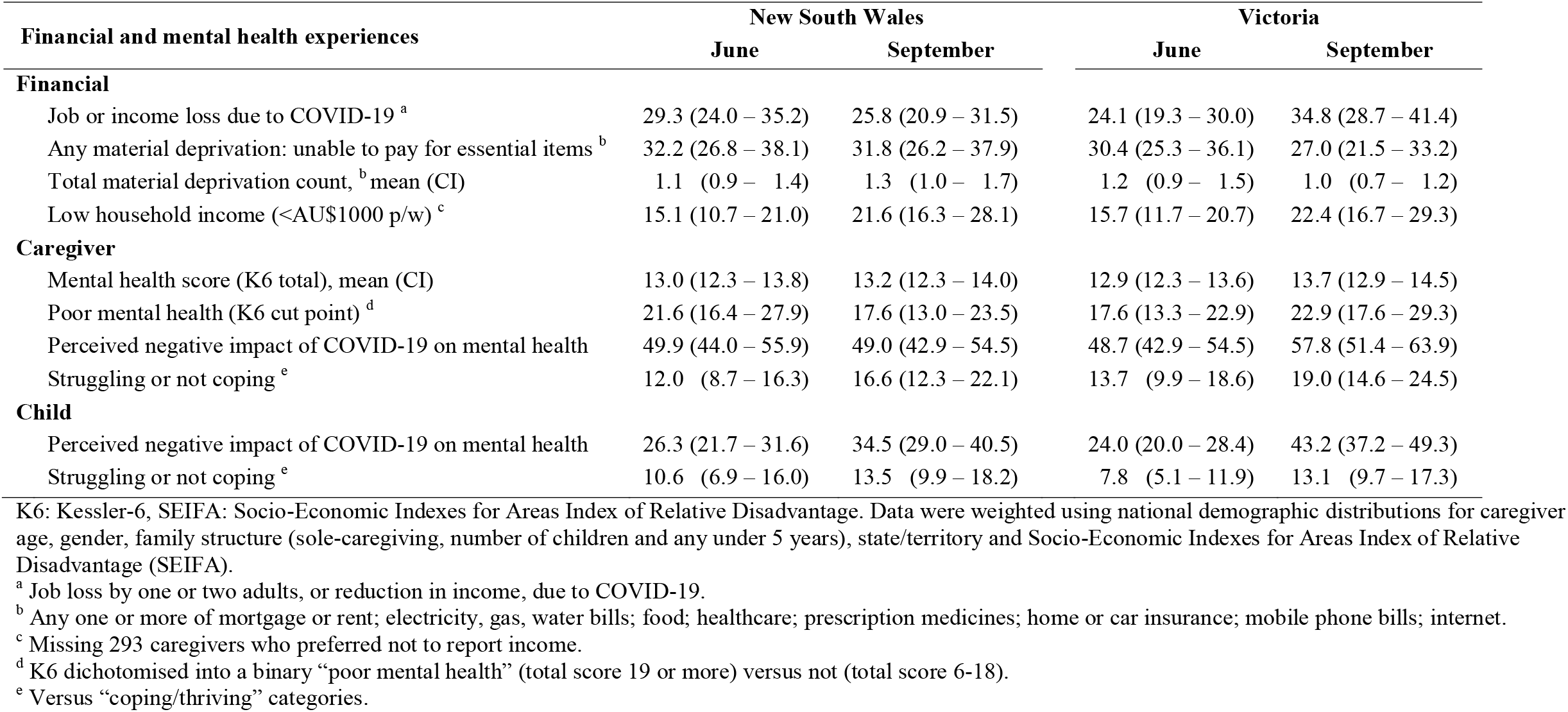
Financial and mental health experiences (weighted) of New South Wales and Victorian families surveyed after initial lockdown (June 2020) and during Victoria’s ongoing lockdown (September 2020) in proportions (95% confidence intervals) unless specified.

### The experiences of ongoing lockdown (Aim 2)

Table 3 presents the Difference-in-Difference analyses testing the relationships between ongoing lockdown and families’ finances and mental health, for the cohorts overall and by subgroups. To enable interpretation, Figure 1 graphs the binary variables. Over the June-September 2020 period, relative to their NSW counterparts, Victorians reported an 11% increase in job or income loss (95% CI: 3 to 18%). There was no evidence that ongoing lockdown was related to a change in material deprivation or the proportion of households reporting low income. Ongoing lockdown was associated with a mean 0.83 increase (95% CI: -0.08 to 1.74) in K6 total score and 6% increase (95% CI: -0.3 to 12%) in the binary K6 measure of poor mental health. Perceived negative mental health impacts of the pandemic was also associated with ongoing lockdown, reported by 14% (95% CI: 6 to 23%) more Victorian caregivers for themselves and for 12% (95% CI: 4 to 20%) more Victorian children relative to NSW.

**Table 3.**
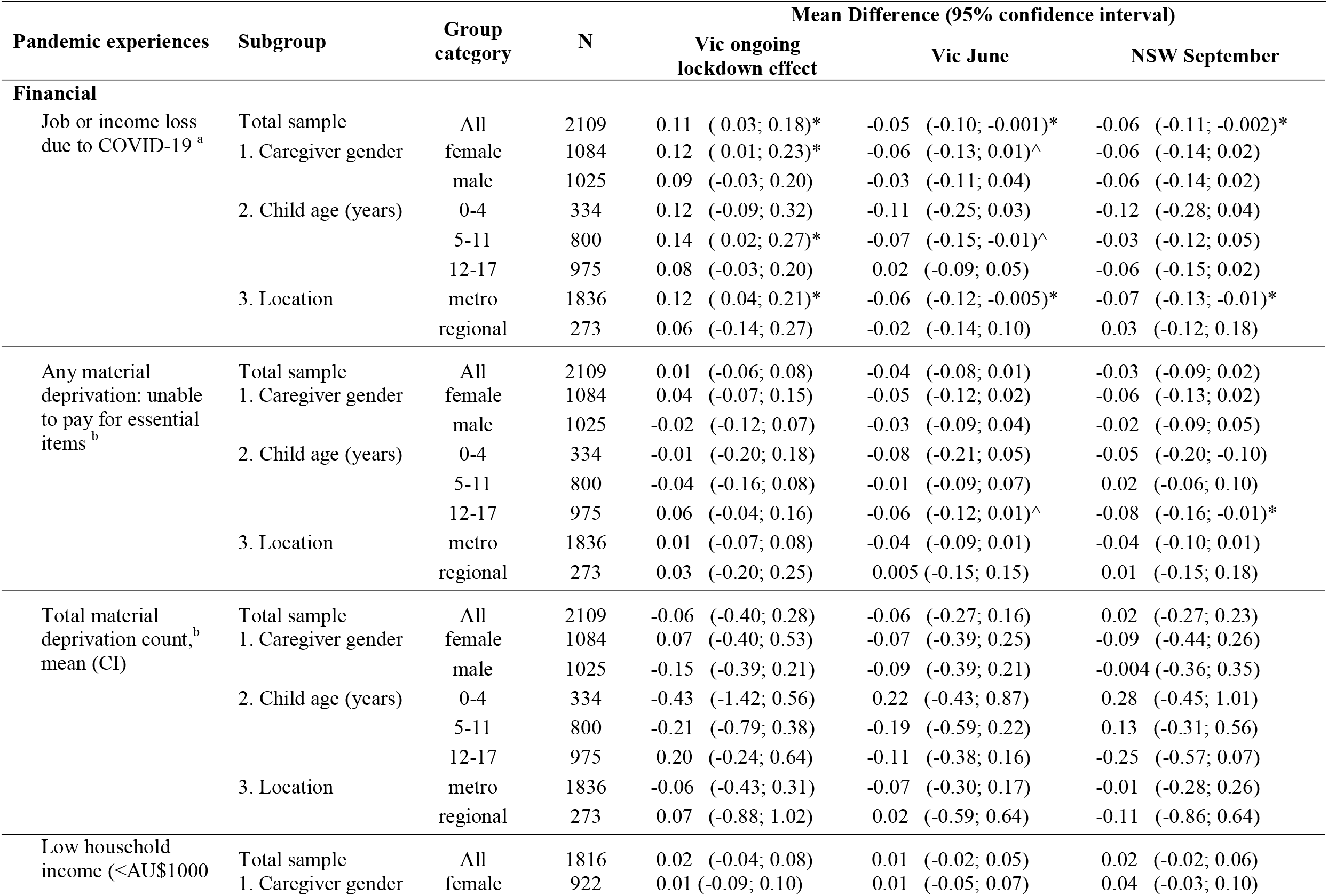
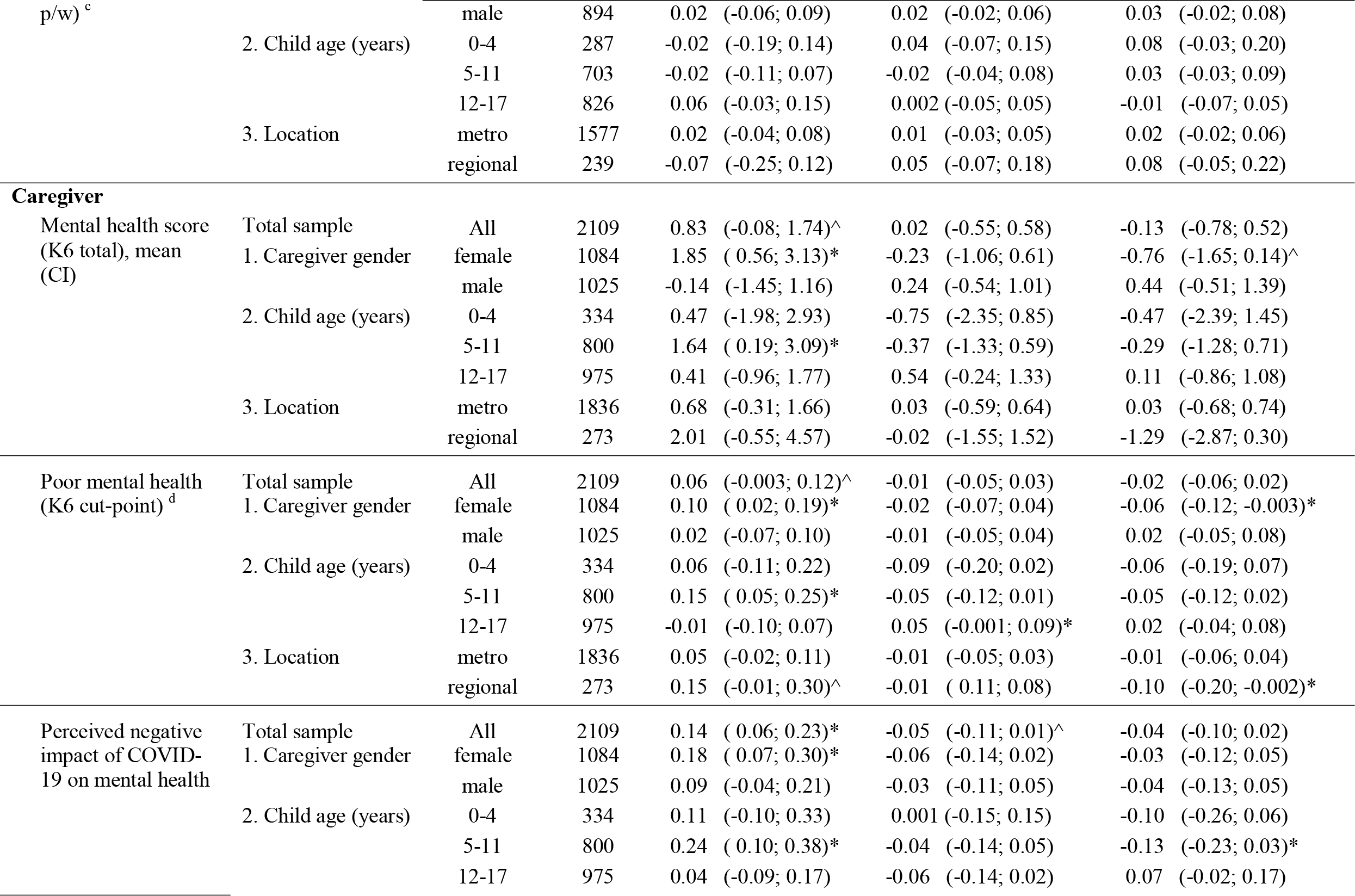
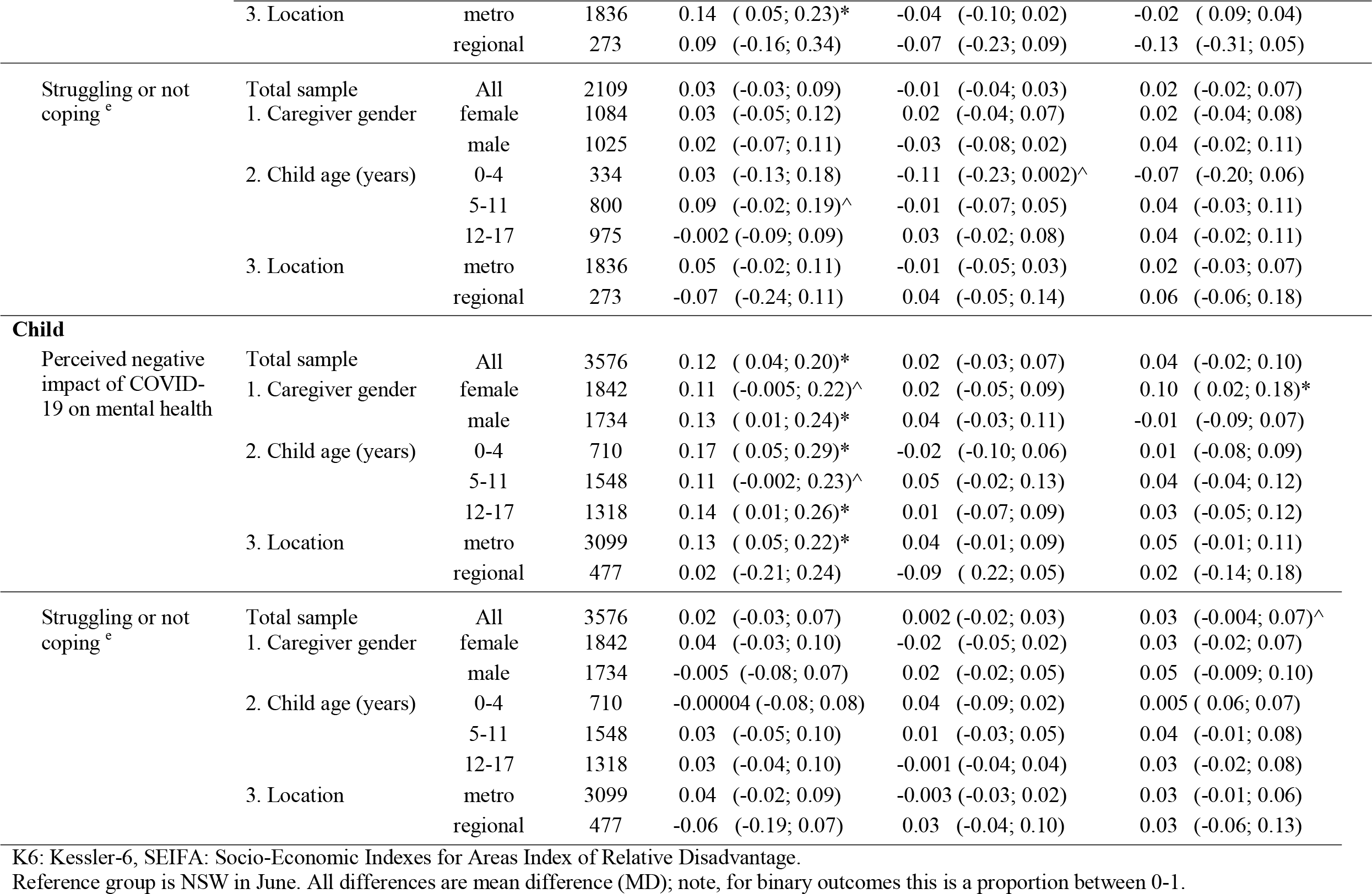
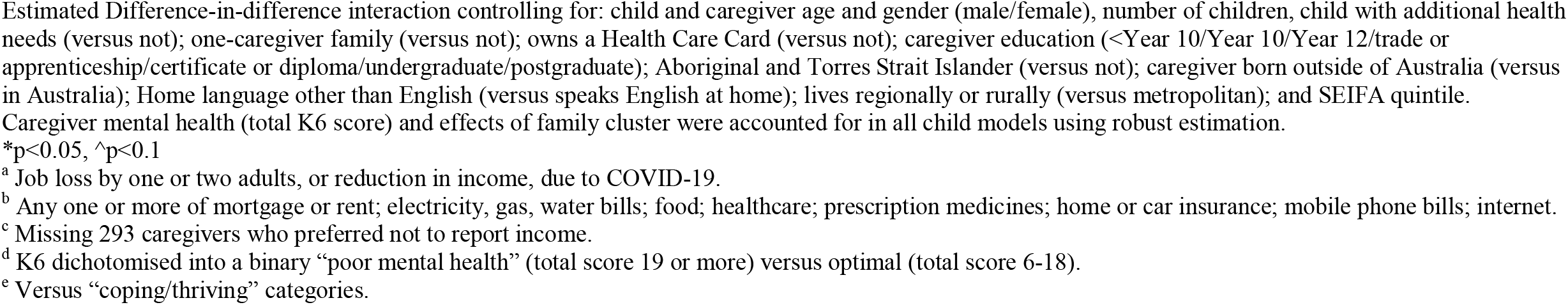
Difference-in-Difference linear regression models estimating the relationship between ongoing lockdown and families’ finances and mental health for the Total sample and by subgroups, using New South Wales (NSW) families in June 2020 as reference group.

**Figure 1.**
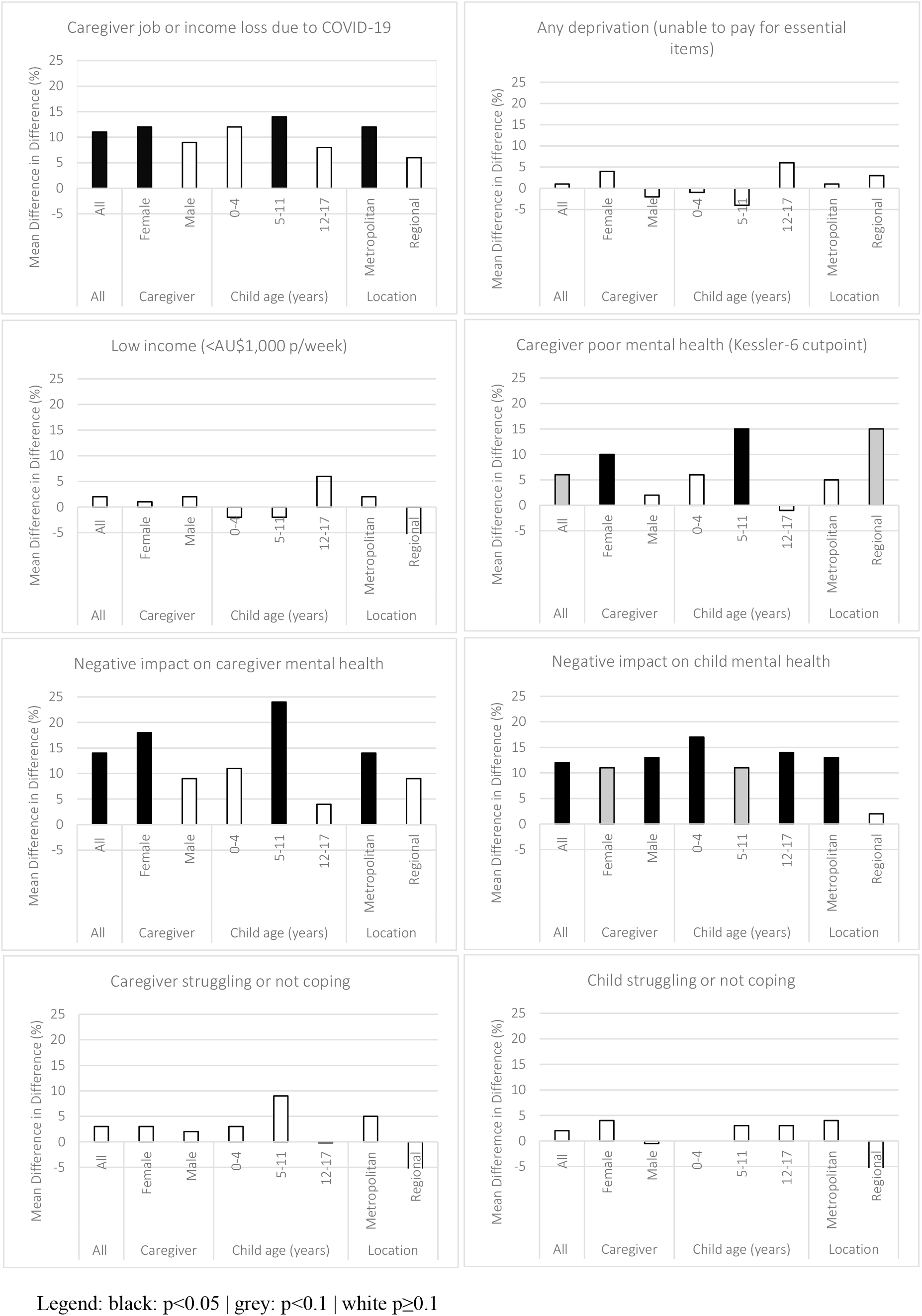
Difference-in-Difference estimates (%) of the relationships between ongoing lockdown and families’ finances and mental health (all binary), overall and by subgroups, data drawn from Table 3.

When considering the subgroups, the evidence for a relationship between ongoing lockdown and job/income loss and caregiver mental health was strongest for female caregivers, caregivers of 5-11-year-old children, and caregivers living in metropolitan areas. Notably, Table 3 (‘NSW September’ column) shows that the differences between initial and ongoing lockdowns in measures of job/income loss and K6 mental health were in part due to worsening outcomes for Victorians as well as improving outcomes for NSW families. The proportion of NSW caregivers reporting job/income loss decreased by 6% (95% CI: 0.2-11%) from June to September. The same reduction was evident in the poor mental health (K6) of female caregivers (mean difference=6%, 95% CI: 0.3 to 12%). There was evidence that ongoing lockdown was related to caregivers’ perception of the pandemic having negative impacts on their children’s mental health, for all subgroups except for families living in regional/rural areas. There was no evidence of a relationship between ongoing lockdown and whether the caregiver or their children were coping or not, for the cohort overall or by subgroups.

## Discussion

This study investigated the relationships between COVID-19 lockdown and family finances and mental health in the context of Australia’s minimal disease burden in the first year of the pandemic. In June 2020, after an initial national lockdown from March-May, a quarter of Australian caregivers of children aged 0-17 years reported job or income loss due to the pandemic. One in three reported material deprivation (being unable to afford essential items such as housing, food, amenities, or healthcare). One in five caregivers reported poor mental health; half said that the first three months of the pandemic had negatively impacted their mental health; and a quarter perceived the same negative impact for their children. By September 2020, Victoria’s ongoing lockdown (from July onwards) was associated with increased job and income loss, and negative mental health impacts for caregivers and children. There was no evidence that ongoing lockdown was related to families’ experiences of material deprivation or income poverty. While the negative experiences of ongoing lockdown were reported by families across children’s ages, they were most pronounced for families with children aged 5-11 years compared with 0-4 or 12-17 years.

The financial and mental health experiences reported by the June cohort are consistent with national data. In April, the ABS estimated that 2.7 million Australians (almost 20% of the working population) lost their jobs or hours of work.(36) The Australian Temperament Project survey of 498 families from March-September 2020,(31) and the right@home trial survey of 319 mothers from May-December 2020,(37) reported job/income losses of 24% and 27% respectively, using the same questions as our study. The third of families reporting material deprivation is equivalent to pre-pandemic data from the Longitudinal Study of Australian Children.(38)

In our study, one in five families reported poor mental health in June 2020 according to the K6. This aligns with the Australian ‘Pulse of the Nation’ survey in which 24% of parents reported mental distress across the first months of lockdown (measured with a single item that highly correlated with the K6).(39) While the RCH Poll lacked pre-pandemic data, substantially more caregivers reported poor mental health on the K6 in the Polls than representative Australian adult data collected pre-pandemic (8% in 2017) or during the first national lockdown (11%).(40) Our study also found a differential increase in the poor mental health of Victorian caregivers of 6.7%. This is significant in statistical and absolute terms, equating to an additional 78,200 parents in Victoria (of 1,167,400) in need of mental health treatment. This finding has significant service implications in terms of ensuring that parents and their children are supported in their increased mental health needs. For example, recent evidence shows the increased mental health presentations by children as result of the pandemic (e.g. a 30-55% increase in presentations to emergency departments of children from socially-advantaged areas in NSW).(41) Despite the global differences between countries’ infection rates and governments’ approaches to public health restrictions, there are commonalities in the mental health data emerging from the pandemic.(42-45) The Born in Bradford study found that 19% and 16% of mothers reported clinically-significant levels of depression and anxiety, respectively, during the first lockdown in UK (April-June 2020).(46) In a nationally weighted survey from the US in June, 27% of parents said their mental health had declined during the pandemic.(47)

Given what is known about the negative economic and psychosocial impacts of lockdown,(42-47) it follows that ongoing lockdown was associated with increased job and income loss, and poor mental health in this study. Our finding that negative mental health experiences were more common for female versus male caregivers is consistent with international and Australian data.(22, 24) That families with children aged 5-11 years were most negatively affected is likely due to the stress and disruption of home-schooling in July-September. More supervision is required for children in elementary/primary school than high school, and balancing home schooling with usual paid or unpaid work was a substantial challenge for families.(48)

Our findings for children are like other studies investigating the lockdown experiences of young people.(29, 49, 50) Analysis by the Australian Human Rights Commission in the earliest months of the pandemic found that increased numbers of older children reported first time mental health challenges or concerns of self-harm.(51) The COVID-19 Unmasked Study which surveyed Australian families with young children (aged 1-5 years) also found that Victorian caregivers exposed to ongoing lockdown reported increasing mental health symptoms for themselves and their children.(52) Interestingly, in the Unmasked study, young children living outside of Victoria (exposed to only the initial lockdown) were still experiencing higher than average levels of anxiety symptoms by the time of Victoria’s ongoing lockdown. We found a similar pattern in our study. For the NSW families exposed to initial lockdown only, caregivers reported improved mental health for themselves by September, but this experience was not evident for their children. It is possible that the shared experience of pandemic stress is a major contributor to the commonalities in international mental health data, over and above the viral incidence or length and severity of lockdown.

This study has several strengths. The large cross-sectional and nationally representative surveys employed strong methodology (piloted and included the validated K6) and achieved high response proportions. In other Polls, indicators (frequency/prevalence) across a range of topics are almost universally consistent with more traditionally obtained estimates, providing support for the sample selection and survey administration methods. The Difference-in-Difference modelling is a well-established method of analysing policy change (in our case, lockdown law differences). However, there are limitations to this analysis. The parallel trends’ assumption supposes that the untreated units (NSW in September) provide the appropriate counterfactual of the trend that the treated units would have followed if they had not been treated. While the time between the first and second surveys was short (four months) – and minimises the potential for differential trends across NSW and Victoria – the lack of pre-pandemic data makes it impossible to verify the validity of this assumption. Further, potential compositional differences in survey participants across time and across states means that there may be other sources contributing to the potential estimated effects.

The reliance on caregiver-report, from only one caregiver per household, means the child rating may be biased by caregiver perception, which is particularly relevant for older children and adolescents. While we controlled for parent mental in the child analyses, we did not collect a validated measure of children’s mental health that would provide a measure of clinical impact. The lack of pre-pandemic data mean the first Poll already captured some of the preliminary effects of the pandemic. However, the intention of the paper was to understand the experiences related to Victoria’s ongoing lockdown, rather than the entire pandemic experience per se. Some caregivers did not disclose family income, and the sample sizes for children aged 0-4 years and regional/rural subgroup analyses were small, limiting power for detecting differences. While we controlled for all available, potentially confounding demographic variables in the analyses, the complex nature of adversity and mental health means that potential unknown residual confounders may have affected the estimates. For example, that the responding cohorts were more advantaged than the general population highlights the importance of research to investigate the experience of syndemic subgroups who are most likely to be negatively affected.(23)

This work extends the evidence base by investigating the indirect and negative experiences of ongoing lockdown in the context of minimal disease burden. We offer three considerations for pandemic response and recovery planning. First, while job and income loss increased with ongoing lockdown, this did not translate to increased material deprivation or income poverty. This finding provides support for the effectiveness of the Australian government’s extraordinary income supplements, introduced early in the pandemic to offset the anticipated economic fallout of lockdown. This interpretation is supported by modelling demonstrating the substantial reductions in Australia’s poverty levels subsequently.(16) Given that the income supplements offered were temporary, financial security must be considered when enacting future lockdowns.

Second, for families who were unexposed to ongoing lockdown, there was some recovery in employment/income and female caregivers’ mental health. While comparable data on financial experiences are limited, the global mental health evidence also shows a recovery for many adults following an initial peak in psychological distress.(20) However, the available systematic reviews are limited by over-representation of data from the early months of the pandemic,(29, 49, 50) and previous pandemics show that negative mental health effects can persist.(52) While the Poll data were limited, they did not suggest a recovery for children. Thus, it is important that children’s experiences and needs are prioritised during response and recovery planning so that any persistent negative impacts are adequately redressed.(54, 55)

Third, the negative financial and mental health experiences related to ongoing lockdown were substantial, and disproportionately affected families with children aged 5-11-years (corresponding with elementary/primary school), and female caregivers. While this study was underpowered to investigate the experience of families living in lower socioeconomic environments, the evidence suggests that inequity is likely to be exacerbated and entrenched by the social and economic disruption of COVID-19.(10, 32, 55, 56) Ongoing follow-up of cohorts is necessary to understand if and how caregivers and children recover from lockdown, and how best to support the population groups who are most adversely affected.

Balancing the benefits and harms of lockdown requires planned responses to future outbreaks, and evidence-informed financial and mental health supports. The clear nexus between the pandemic and inequitable associations with poorer mental health suggests a need to respond now through policy-focussed action on mental health prevention (including financial support) and to plan for future lockdowns through evidence-informed financial and direct mental health supports.

## Data Availability

The data underlying the results presented in the study are available from the RCH
Child Health Poll, please contact.

## Acknowledgments

We thank all families who took part in the Royal Children’s Hospital National Child Health Polls. We thank Monsurul Hoq, the statistician for the RCH National Child Health Poll, for preparing the data for this study. The National Child Health Polls are supported by The Royal Children’s Hospital Foundation. This research was supported by the Helen Macpherson Smith Trust Impact Grant #9523. The Murdoch Children’s Research Institute (MCRI) administered the research grants for the work and provided infrastructural support (as study sponsor) to its staff but played no role in the conduct or analysis of the research. Research at the MCRI is supported by the Victorian Government’s Operational Infrastructure Support Program. New South Wales authors were supported by the Population Child Health Group at the University of New South Wales, and Best-START South West at the Ingham Institute. Author AP was supported by the Erdi Foundation Child Health Equity Scholarship and The Corella Fund. Author SW was supported by a National Health and Medical Research Council (NHMRC) Career Development Fellowship (#1158954); Author SG was supported by a NHMRC Practitioner Fellowship (#1155290).

## Supplementary information

**Supp. Figure 1. Timeline of Australia’s COVID-19 public health restrictions and policy changes from March to November 2020**

**Supp. Figure 2. Respondent flowchart for analytic sample**

