## Supplementary Fig for "The relationships between ongoing COVID-19 lockdown and the financial and mental health experiences of Australian families"

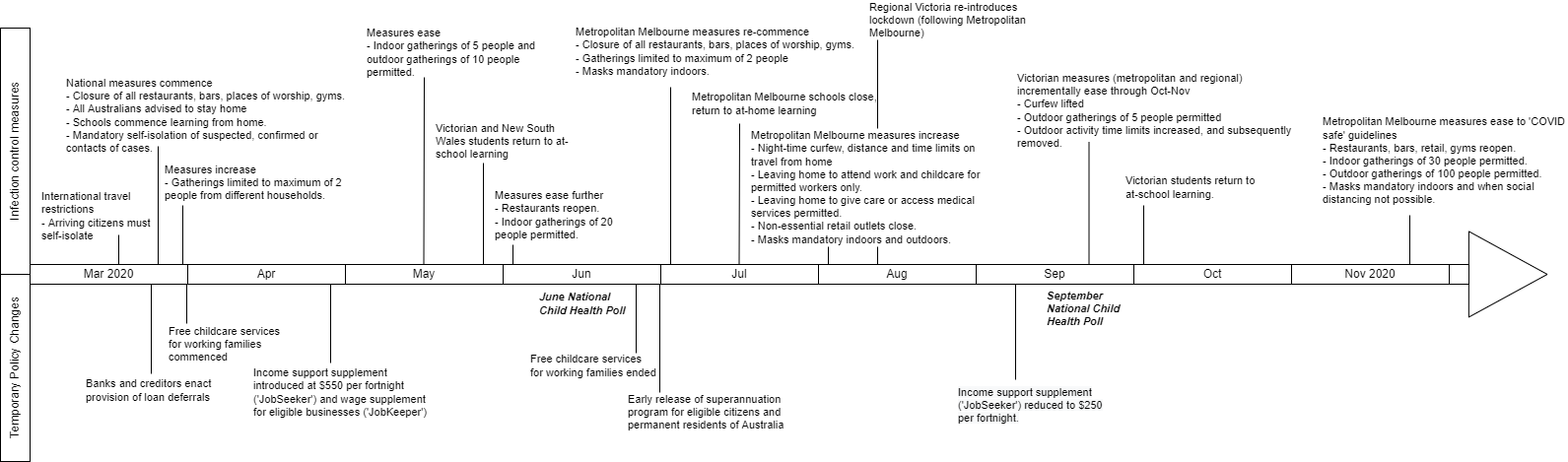

**S1 Fig. Timeline of Australia’s COVID-19 public health restrictions and policy changes from March to November 2020.**

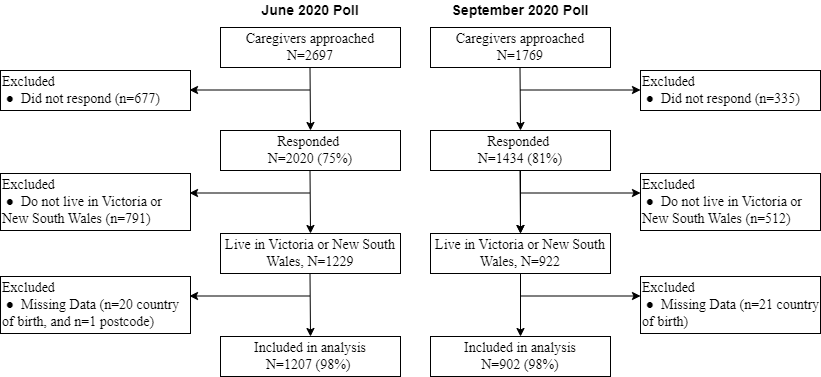

**S2 Fig. Respondent flowchart for analytic sample.**
